# Real-time breath metabolomics to assess early response to CFTR modulators in adults with cystic fibrosis: an open-label proof-of-concept study

**DOI:** 10.1101/2024.05.29.24308131

**Authors:** Emmanuelle Bardin, Hélène Salvator, Camille Roquencourt, Elodie Lamy, Nicolas Hunzinger, Isabelle Sermet-Gaudelus, Sandra De Miranda, Dominique Grenet, Philippe Devillier, Stanislas Grassin-Delyle

**Affiliations:** Université Paris Cité, INSERM U1151, CNRS UMR8253, Institut Necker Enfants Malades, Paris, France; Université Paris-Saclay, UVSQ, INSERM, Infection et inflammation (2I), U1173, Département de Biotechnologie de la Santé, Unité Technologies pour la Santé et le Médicament, Montigny le Bretonneux, France; Hôpital Foch, Exhalomics®, Suresnes, France; Hôpital Necker Enfants Malades, Centre de Référence Maladies Rares, Paris, France; Université Paris-Saclay, UVSQ, UFR Simone Veil-Santé, VIM-Suresnes, UMR0892, Suresnes, France; Hôpital Foch, Service de pneumologie, CRCM - Centre de Transplantation Pulmonaire, Suresnes, France

**Keywords:** Breathomics, cystic fibrosis, CFTR modulators, real-time mass spectrometry, volatile organic compounds

## Abstract

**Background:** Highly effective CFTR modulators, particularly elexacaftor/tezacaftor/ivacaftor (ETI), produce rapid clinical improvements in people with cystic fibrosis. Yet early treatment effects may be difficult to capture with spirometry or sweat test in patients with a mild disease or atypical mutations. Exhaled breath is rich in volatile organic compounds (VOCs) reflecting metabolic and inflammatory processes. We aimed to determine whether ETI induces early, measurable changes in breath composition and whether these changes relate to clinical outcomes.

**Methods:** Ten adults initiating ETI were enrolled in a prospective, open-label study with breath sampling at baseline, week one and month one. VOCs were measured using real-time proton-transfer-reaction - mass spectrometry (PTR-MS). Longitudinal changes were assessed using multilevel statistics, including univariate linear mixed-effects models and multivariate RM-ASCA+; repeated-measures correlations examined associations with lung function and sweat chloride concentration. Results were compared with a healthy cohort.

**Results:** Amongst the eight responders, 11 VOC features changed significantly after ETI initiation. Eight differed from healthy controls at baseline and shifted towards healthy levels over one month. RM-ASCA+ identified monotonous and non-monotonous patterns capturing various dynamics such as acute, progressive or delayed metabolic responses. A 11-feature PLS-DA model classified visits with high accuracy (AUC=0.84-0.96). Ten VOCs correlated with clinical readouts. Features of interest were tentatively identified and pointed towards a shift in the microbiome and/or energy metabolism.

**Conclusions:** ETI induces rapid alterations in exhaled VOCs, many trending towards healthy values and correlating with clinical improvement. Real-time breath analysis offers a promising non-invasive surrogate for early monitoring of therapeutic response.

**HIGHLIGHTS:**

- ETI induces rapid, measurable changes in exhaled volatile organic compounds.
- Breath VOCs shift towards healthy levels within one week of ETI initiation.
- Real-time PTR-MS enables non-invasive, point-of-care breath monitoring in CF.
- Multilevel statistics identify monotonous and non-monotonous VOC trajectories.
- Ten VOCs correlate with lung function and sweat chloride.tab

## 1 Introduction

The development of cystic fibrosis transmembrane conductance regulator (CFTR) modulators is transforming the therapeutic landscape for patients with cystic fibrosis (CF). In eligible patients, the combination of the potentiator ivacaftor with the two correctors tezacaftor and elexacaftor (ETI) achieves unprecedented improvements in lung function and quality of life within a few weeks [1]. Correctors support the intra-cellular processing of misfolded CFTR proteins, produced by class II mutations such as the most common p.Phe508Del (F508del thereafter), whilst the potentiator improves the functioning of CFTR channels positioned at the apical side of epithelial cells. In pulmonary cells, the restoration of CFTR activity reinstates the ionic osmotic balance, which translates into rehydration of lung-lining fluids and restoration of mucociliary clearance [2]. ETI is indicated for patients with at least one non-Class I mutation that produces a CFTR protein amenable to correction.

However, these highly efficient therapies came with poorly anticipated challenges. Clinical evaluation of people with CF (pwCF) mainly focuses on respiratory function through the monitoring of the forced expiratory volume in one second (FEV_1_) and the forced vital capacity (FVC), since the respiratory status still mostly determines prognosis. Yet, these endpoints sometimes fail to detect clinical benefit or biological changes, for example in pwCF with high baseline lung function and in young children unable to perform these tests or, conversely, in people with irreversible lung damage [3,4]. The measurement of sweat chloride concentration (SCC) often serves as a secondary endpoint in clinical trials as a marker of CFTR activity, although it does not always correlate with clinical improvement, e.g. in respiratory function and symptoms [5,6]. Some patients show delayed or limited effects on lung function or SCC [5,7], making these outcomes unreliable for assessing drug efficacy in certain cases. Nevertheless, other clinical benefits are emerging, such as reduced exacerbation frequency or lower bacterial load [8], normalisation of inflammation [9], or improved nutritional status [10], although long-term data are still lacking on the newest CFTR modulators combinations. Therefore, there is a rationale for developing alternative methods capable of capturing early therapeutic effects of novel treatments across patients with diverse mutations and varying disease severity.

Over the past fifty years, breathomics has expanded across multiple domains of respiratory medicine and has emerged as a promising, innovative approach. Exhaled breath constitutes a unique, individual-specific biological signature, rich in personalised metabolic information, sensitive to subtle physiological changes, and obtainable entirely non-invasively [11]. Exhaled metabolites (volatile organic compounds (VOCs)), are believed to arise from inflammatory processes, oxidative reactions, and enzyme-driven metabolic pathways, all of which may be modulated by pharmacological treatments. VOCs have been associated with a wide range of conditions, including chronic and acute pathologies, infections, environmental exposures, and therapeutic interventions [12]. Notably, a previous study following up 20 F508del-homozygous patients initiating the CFTR modulator dual therapy lumacaftor/ivacaftor reported significant metabolic changes in exhaled breath profiles after three months which persisted at 12 months [13]. Online proton-transfer reaction - mass spectrometry (PTR-MS) is a high-sensitivity, point-of-care technology allowing real-time monitoring of VOCs which has already demonstrated potential for medical diagnosis [14–16]. We hypothesised that ETI-induced metabolic modifications could be detected in exhaled breath using PTR-MS, and that alterations in volatilomic profiles might be associated with clinical outcomes.

## 2 Materials and Methods

### 2.1 Study design

We conducted a prospective, open-label study at the Foch university hospital (Suresnes, France), enrolling any adult with CF (≥18 years) initiating ETI. Patients were participating to the PHEAL-KAFTRIO study, approved by an ethics committee (Comité de protection des personnes Sud-Est I, 2021-A03119-32, NCT05295524). All patients provided written informed consent. The study included three visits: before ETI initiation (V0), during the first week (V1) and after one month (V2) of treatment. At each visit, clinical readouts including respiratory symptoms (cough, sputum production, dyspnoea) and episodes of exacerbation were recorded. Spirometry outcomes (FEV_1_ and FVC in absolute and percentage of predictive values (pp)) were measured according to ATS/ERS guidelines [17]. Sweat collection was performed using a Macroduct^®^ system according to the recommendations of the ECFS [18]. As established in previous reference publications [5], response to ETI was defined by functional criteria: +10% ppFEV_1_ and/or a drop of more than 20 mmol/L in SCC. The response of patients with compassionate access to ETI, i.e. carrying mutations excluded from the initial indication, were evaluated after two months of treatment by an adjudication committee [5]. The data were compared with those obtained using the same instrument in a cohort of healthy young adults (VOC-COMPARE study, NCT06020521, 40 individuals with matching age enrolled between July-August 2023 [19]).

### 2.2 Breath analysis

At each visit, breath volatile metabolites were analysed with real-time mass spectrometry using a proton-transfer reaction – quadrupole interface – time-of-flight mass spectrometer (PTR-Qi-TOF MS; Ionicon, Innsbruck, Austria). Patients blew directly into the instrument via a single-use mouthpiece. Five exhalations of ∼15 s with 40 s intervals were performed (<5 min total), requiring minimal patient cooperation. Operating parameters are detailed in the Supplementary Methods.

### 2.3 Breath analysis data processing

Data were processed following a workflow detailed in the Supplementary Methods, including mass calibration, exhalation phase detection, peak deconvolution (Supplementary Figure 1), quantification, alignment, background filtering, and missing value imputation. Values were expressed as counts per second (cps) after probabilistic quotient normalisation (PQN), log-transformation and autoscaling. Putative annotations were proposed based on available databases and literature and an in-house VOC library (Supplementary Methods).

### 2.4 Statistical analysis

The effect of treatment was studied in responders; correlations were assessed in all patients. The full statistical workflow is shown in Supplementary Figure 2 and detailed in Supplementary Methods. Longitudinal changes were assessed using a linear mixed-effects model (LMM) for each feature, with visit as a fixed effect and patient as a random effect. A repeated measures ANOVA– simultaneous component analysis plus (RM-ASCA+) was then applied to visualise main temporal patterns and quantify the proportion of variance explained by time. A multilevel PLS-DA model was built with leave-one-out cross-validation at the patient level. A recursive feature elimination (RFE) algorithm was applied to identify the optimal feature subset. Comparisons with healthy adults used Wilcoxon-Mann-Whitney tests; correlations with clinical readouts used a repeated measure correlation test [20]. All *p*-values were adjusted using the Benjamini–Hochberg FDR correction.

## 3 Results

### 3.1 Study population and clinical response

Ten participants (two females), median age 34 years (interquartile range (IQR) 17), were enrolled from March to December 2022; three had compassionate access and two switched from monotherapy ivacaftor (Table 1). At baseline, lung disease stage was variable; all patients were positive for at least one respiratory pathogen (*Pseudomonas aeruginosa* (5/10), *Staphylococcus aureus* (5/10)). Seven had daily anti-infectious prophylaxis (inhaled and oral route) throughout the study (Supplementary Table 1). Over the course of the study, three patients experienced a mild bronchial exacerbation; two of them were related to a coronavirus disease (COVID-19) infection and required additional antibiotics (Supplementary Table 1).

**Table 1.** Cohort characteristics and clinical readouts during the study.

| Patient |  | 01 | 02 | 03 | 04 | 05 | 06 | 07 | 08 | 09 | 10 |
| --- | --- | --- | --- | --- | --- | --- | --- | --- | --- | --- | --- |
| Sex |  | F | M | M | M | M | M | M | M | F | M |
| Age range (y) |  | 26-30 | 41-45 | 36-40 | 31-35 | 51-55 | 31-35 | 21-25 | 46-50 | 21-25 | 66-70 |
| Genotype |  | F508del/<br>S1255P | F508del/<br>F508del | F508del/<br>F508del | F508del/<br>S945L | S945L(exon15)<br>/ I601F | 3120G>A/<br>3120G>A | N1303K/<br>N1303K | F508del/<br>E92K | F508del/<br>I148T | F508del/<br>3849+10kbC>T |
| CFTR<br>modulator<br>treatment at V0 |  | Ivacaftor | - | - | - | Ivacaftor | - | - | - | - | - |
| Therapy access |  | Eligible | Eligible | Eligible | Eligible | Off-label | Off-label | Off-label | Eligible | Eligible | Eligible |
| SCC<br>(mmol/L) | V0 | 15 | Failed | 92 | 100 | 24.5 | 106 | 96 | 92.5 | 100 | 64 |
|  | V1 | <10 | ND | 17 | 60 | 28.5 | 124 | 93 | 38 | 51.5 | 32.5 |
|  | V2 | ND | ND | ND | 58.5 | <10 | 100.5 | 108 | 21.5 | 52 | 44 |
| ppFEV<sub>1</sub><br>(%) | V0 | 35 | 54 | 120 | 80 | 24 | 30 | 18 | 67 | 87 | 76 |
|  | V1 | 38 | 66 | 129 | 91 | 25 | 28 | 20 | 74 | 96 | 78 |
|  | V2 | 43 | 80 | 137 | 94 | 25 | 28 | 21 | 77 | 96 | 87 |
| ppFVC<br>(%) | V0 | 64 | 61 | 125 | 89 | 63 | 48 | 55 | 88 | 99 | 80 |
|  | V1 | 68 | 94 | 128 | 99 | 51 | 46 | 61 | 91 | 103 | 81 |
|  | V2 | 71 | 103 | 124 | 100 | 52 | 46 | 72 | 94 | 105 | 91 |
| Clinical<br>improvement<br>at V2 |  |  | Cough<br>Dyspnea<br>+3kg | Cough<br>Sputum | Cough<br>Sputum<br>+5kg | Fatigue<br>Sputum<br>+1kg |  | Dyspnea<br>Sputum<br>+1kg | Cough<br>Dyspnea<br>Sputum | Cough<br>Dyspnea<br>Sputum | Cough<br>Dyspnea<br>+2kg |
| Response to ETI |  | Inconclusive | Yes | Yes | Yes | Yes | No | Yes* | Yes | Yes | Yes |
| ETI<br>interruption |  | at one<br>month | No | No | No | No | at two<br>months | No | No | No | No |

V0: baseline; V1: week 1; V2: month 1 of elexacaftor/tezacaftor/ivacaftor (ETI) treatment. SCC: sweat chloride concentration; ppFEV<sub>1</sub>: percent predicted in forced expiratory volume in 1 second; ppFVC: percent predicted in forced vital capacity; ND = Not done. \*as evaluated at two months treatment according to the compassionate programme protocol [5]

At V2, six patients carrying F508del experienced clinical improvement and met clinical response criteria (Table 1). One patient switching from ivacaftor chose to stop ETI after one month because of insomnia; despite a +8% gain in ppFEV_1_ his response was deemed inconclusive. Amongst the three patients who were granted off-label ETI access, one exhibited a quick response, the second showed a delayed response with +10% ppFEV_1_ after two months, and the third discontinued the treatment based on poor clinical outcomes. This resulted in a total number of 8/10 responders. Side effects were limited to rash (3/10) and insomnia (1/10). Microbiological cultures in patients able to cough up sputum (8/10) were qualitatively unchanged compared to those collected at baseline.

### 3.2 Breath analysis

#### 3.2.1 Detection of VOCs

A total of 29 breath samples were obtained, from which 99 features were consistently detected. Five groups of correlated features were identified, corresponding to water clusters or dehydrated forms of alcohols or short chain fatty acids (SCFA): *m/z* [33.03/51.04] (methanol and water cluster); *m/z* [47.05/65.06] (ethanol and water cluster); *m/z* [61.03/79.04/43.02] (acetic acid; ±H_2_O); *m/z* [75.04/57.03] (propionic acid; -H_2_O); *m/z* [89.06/71.05/107.08] (butyric acid; ±H_2_O). Redundant information from these groups was eliminated by keeping only parent ions, which resulted in a total of 92 features available for statistical analysis (Supplementary Table 2). Unsupervised PCA demonstrated a homogeneous dataset, with patient identity representing the dominant source of variability (Supplementary Figure 3).

#### 3.2.2 Changes in breath profiles

Longitudinal analysis using LMM identified 11 features that changed in responders following ETI initiation (*p*-value <0.05, Table 2, Figure 1). Of these, eight exhibited different levels in pwCF at V0 compared to healthy volunteers (*p*-value <0.05). Modifications in CF breath profiles were detectable from V1 onwards, progressively shifting towards healthy levels. By V2, seven features remained differentially expressed in pwCF.

**Table 2.** VOCs significantly modified upon ETI treatment based on LMM or PLS-DA models, and/or correlated with variations in clinical readouts.

| *m/z* | Exhaled<br>breath<br>(mean,<br>ppb) | LMM model |  |  | Selected<br>with the<br>PLS-DA<br>model | Correlations |  |  |  |  |  | Molecular<br>formula | Tentative annotations<sup>#</sup> |
| --- | --- | --- | --- | --- | --- | --- | --- | --- | --- | --- | --- | --- | --- |
| | | *p*-<br>value | $\beta_1$<br>(V1-V0) | $\beta_2$<br>(V2-V0) | | ppFEV<sub>1</sub> | | ppFVC | | SCC | | | |
|  |  |  |  |  |  | *p*-value | R | *p*-value | R | *p*-value | R |  |  |
| <u>33.03</u>/ 51.04 | 395.33 |  |  |  | Yes |  |  |  |  |  |  | [CH<sub>4</sub>O+H]<sup>+</sup> | Methanol cluster |
| 41.04 | 246.94 |  |  |  | Yes |  |  |  |  |  |  | [C<sub>3</sub>H<sub>4</sub>+H]<sup>+</sup> | Fragments |
| <u>47.05</u>/ 65.06 | 146.45 |  |  |  | Yes |  |  |  |  |  |  | [C<sub>2</sub>H<sub>6</sub>O+H]<sup>+</sup> | Ethanol cluster |
| 55.05 | 7.86 | 0.041 | -0.22 | -0.06 | Yes |  |  |  |  |  |  |  | Fragments |
| 57.07 | 9.47 | 0.029 | 1.16 | 1.22 | Yes |  |  | 0.050 | 0.49 |  |  | [C<sub>4</sub>H<sub>8</sub>+H]<sup>+</sup> | C4-Alkenes and fragments |
| <u>61.03</u>/ 43.02/<br>79.04 | 32.64 | 0.052 | -0.32 | -0.17 |  |  |  |  |  |  |  |  | Acetic acid cluster |
| 63.04 | 1.07 | 0.027 | 0.10 | -0.32 | Yes |  |  |  |  |  |  |  | Acetaldehyde+H<sub>2</sub>O |
| 67.07 | 0.55 | 0.016 | -0.57 | -1.11 |  |  |  |  |  |  |  |  |  |
| 69.09 | 20.93 |  |  |  |  |  |  |  |  | 0.011 | 0.62 | [C<sub>5</sub>H<sub>8</sub>+H]<sup>+</sup> | Isoprene |
| 71.09 | 2.41 |  |  |  | Yes |  |  | 0.004 | -0.63 |  |  |  | C5-Alkenes and fragments |
| 73.06 | 1.73 |  |  |  |  |  |  | 0.015 | -0.65 |  |  |  | 2-Butanone |
| 74.06 | 0.31 | 0.038 | -0.15 | -0.63 |  | 0.029 | -0.6 |  |  |  |  | [C<sub>3</sub>H<sub>7</sub>NO+H]<sup>+</sup> | N,N-dimethylformamide,<br>propanamide,<br>propanenitrile+H<sub>2</sub>O |
| 79.08 | 0.38 |  |  |  | Yes |  |  |  |  |  |  |  |  |
| 85.10 | 1.03 | 0.001 | 0.47 | 1.06 | Yes | 0.004 | 0.68 | 0.038 | 0.5 |  |  | [C<sub>6</sub>H<sub>12</sub>+H]<sup>+</sup> | C6-Alkenes and fragments |
| 87.08 | 0.67 | 0.006 | -0.96 | -0.23 | Yes |  |  |  |  |  |  | [C<sub>5</sub>H<sub>10</sub>O+H]<sup>+</sup> | 2-Pentanone |
| <u>89.06</u>/ 71.05/<br>107.08 | 5.38 | 0.051 | -0.27 | -0.33 |  | 0.024 | -0.5 | 0.004 | -0.62 | 0.035 | 0.54 | [C<sub>4</sub>H<sub>8</sub>O<sub>2</sub>+H]<sup>+</sup> | Butyric acid cluster |
| 99.12 | 0.21 |  |  |  |  | 0.037 | 0.49 |  |  |  |  | [C<sub>7</sub>H<sub>14</sub>+H]<sup>+</sup> | C7-Alkenes and fragments |
| 103.08 | 0.19 | 0.053 | -0.12 | -0.33 |  | 0.009 | -0.58 |  |  |  |  | [C<sub>5</sub>H<sub>10</sub>O<sub>2</sub>+H]<sup>+</sup> | Pentanoic acid |
| 135.11 | 0.17 |  |  |  |  |  |  | 0.043 | 0.48 |  |  | [C<sub>10</sub>H<sub>14</sub>+H]<sup>+</sup> | o-Cymene |
| 139.15 | 0.06 | 0.027 | 0.52 | -0.88 | Yes |  |  |  |  |  |  | [C<sub>10</sub>H<sub>18</sub>+H]<sup>+</sup> | C10-Alkenes |

Concentrations in ppb are reported for indicative purposes for the most abundant ion (underlined *m/z* in the case of a cluster); *p*-values obtained with a linear mixed-effects model (LMM) built with patients responding to the treatment elexacaftor/tezacaftor/ivacaftor (ETI); $\beta_x$ LMM coefficients indicates the direction of the evolution at visits V1 and V2 compared to V0; PLS-DA: partial least-squares – discriminant analysis; coefficients (R) of correlation tests (rmcorr) with percent predicted forced expiratory volume in one second (ppFEV<sub>1</sub>), percent predicted forced vital capacity (ppFVC) and sweat chloride concentration (SCC) are provided where significant. <sup>#</sup>Tentative chemical identification is based on exact mass match with the human breathomics database, likelihood of compound ionisation by proton transfer reaction, others' published work and an in-house VOC library (all references available in Supplementary Information). All *p*-values are adjusted with the false discovery rate (FDR) correction.

**Figure 1.**
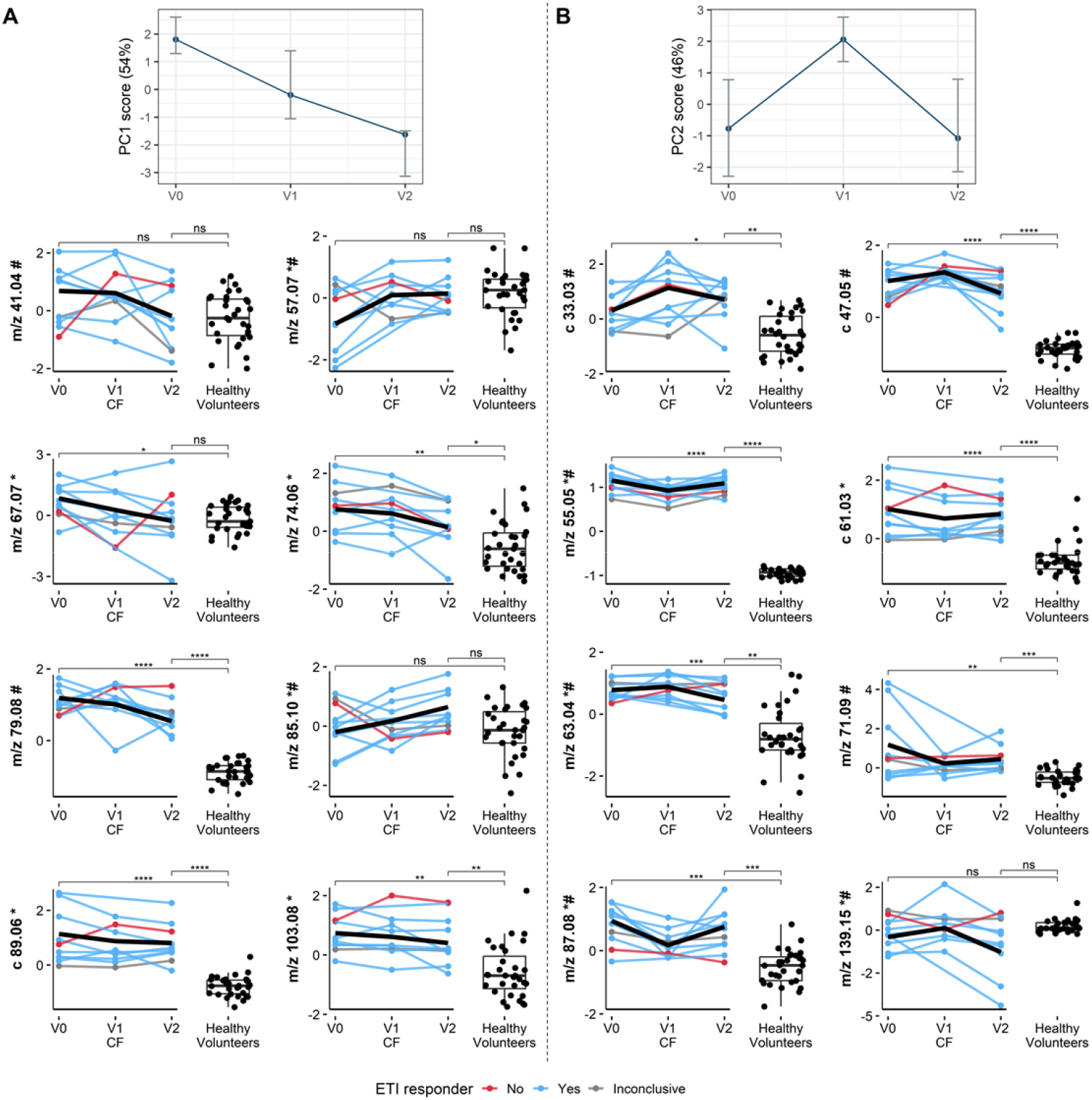
The top two graphs represent the two first principal components (PCs) of the repeated measures ANOVA–simultaneous component analysis plus (RM-ASCA+) model, with 95% percentile bootstrap confidence intervals. The lower plots show the longitudinal evolutions of 16 features in normalised intensities, in the breath of responder patients after elexacaftor/tezacaftor/ivacaftor (ETI) initiation from baseline (V0), to week one (V1) and month one of treatment (V2), compared to levels measured in healthy volunteers. These VOCs correspond to the union of the 11 linear mixed-effects model (LMM) significant features (marked *m/z* *; F-test *p*-value <0.05) and the 11 features selected using recursive feature elimination (RFE) applied to the multilevel partial least-squares – discriminant analysis (PLS-DA) model (marked *m/z* #). The black lines correspond to the fixed effects estimated by the LMM model. Left panel **(A**) shows features most contributing to PC1 of the RM-ASCA+ model; right panel (**B**) shows features most contributing to PC2. \**p*-value <0.05; \*\**p*-value <0.01; \*\*\**p*-value <0.001; \*\*\*\**p*-value <0.0001.

The RM-ASCA+ model indicated that 5% of the total variance was attributable to time. Scores of the first two principal components (PC) are shown at the top of Figure 1 (with corresponding loadings shown in Supplementary Figure 4). PC1, which captures 53% of the time-related variance, reflects features displaying monotonous trajectories over time, either increasing (e.g. *m/z* 85.10) or decreasing (e.g. *m/z* 74.06). In contrast, PC2 (47%) encompasses features with divergent temporal patterns at V1 and V2. It therefore includes a range of profiles, such as features that shifted at V1 and subsequently returned to baseline (e.g. *m/z* 87.08) or stabilised at V2 (e.g. *m/z* 61.03), as well as features that exhibited minimal change at V1 but substantial modifications at V2 (e.g. *m/z* 63.04 and 139.15).

The multilevel PLS-DA model predicted patient visits with an AUC of 0.89 for V0, 0.90 for V1, and 0.72 for V2 without feature selection (Figure 2A). Based on the mean AUC across visits, the RFE method identified an optimal subset of 11 variables (Supplementary Figure 5). Training the model using these 11 features slightly shifted the mean prediction AUC from 0.84 to 0.88. Classification performance for V2 was improved substantially, reaching an AUC of 0.96, whilst V0 and V1 AUCs slightly decreased to 0.84 and 0.86, respectively (Figure 2B). This suggests that the selected features most discriminate V2 compared to the other visits, potentially paralleling therapeutic response. The VIP scores of the 11 selected features are shown in Supplementary Figure 6 and their longitudinal evolution is presented in Figure 1. These overlap with six significant LMM features.

**Figure 2.**
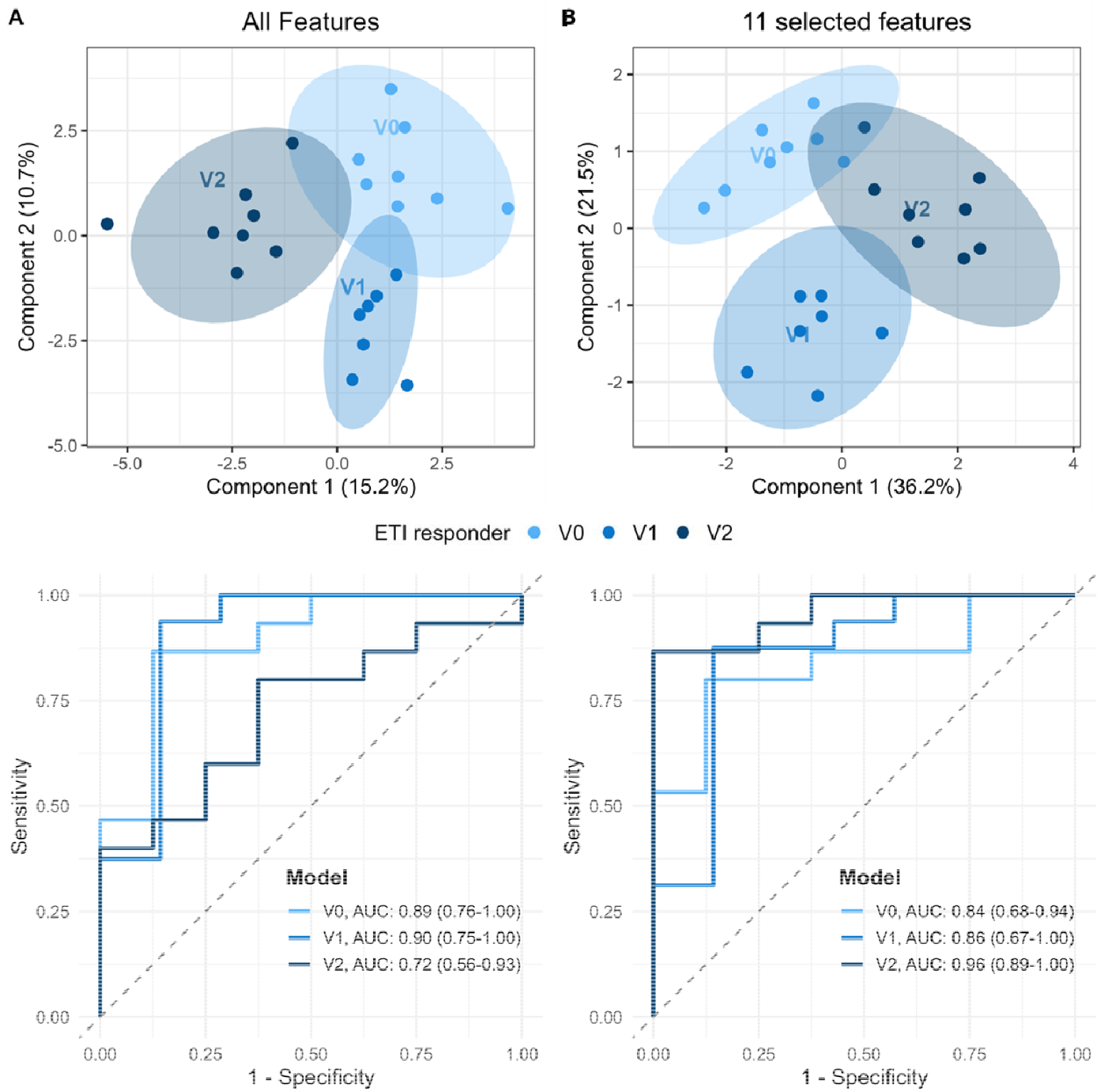
Multilevel partial least-squares – discriminant analysis (PLS-DA) showing the distribution of breath samples collected from responder patients at baseline (V0), after one week (V1), and one month (V2) of elexacaftor/tezacaftor/ivacaftor (ETI) treatment. The top graphs show the scores plots with shaded areas representing 95% confidence intervals, and the bottom graphs show the receiver operating characteristic (ROC) comparing each visit against the others. Left panel **(A)** shows the results of the model using all features (92); right panel (**B**) shows the model built with the 11 features selected with recursive features elimination (RFE).

#### 3.2.3 Correlations with clinical outcomes

A total of 10 breath features were significantly associated with one or more clinical readouts (Table 2, Figure 3, Supplementary Figure 7). Five features correlated with ppFEV□, six with ppFVC, and two with SCC. Notably, five of these ten clinically associated features were also significantly modified following treatment in responders.

**Figure 3.**
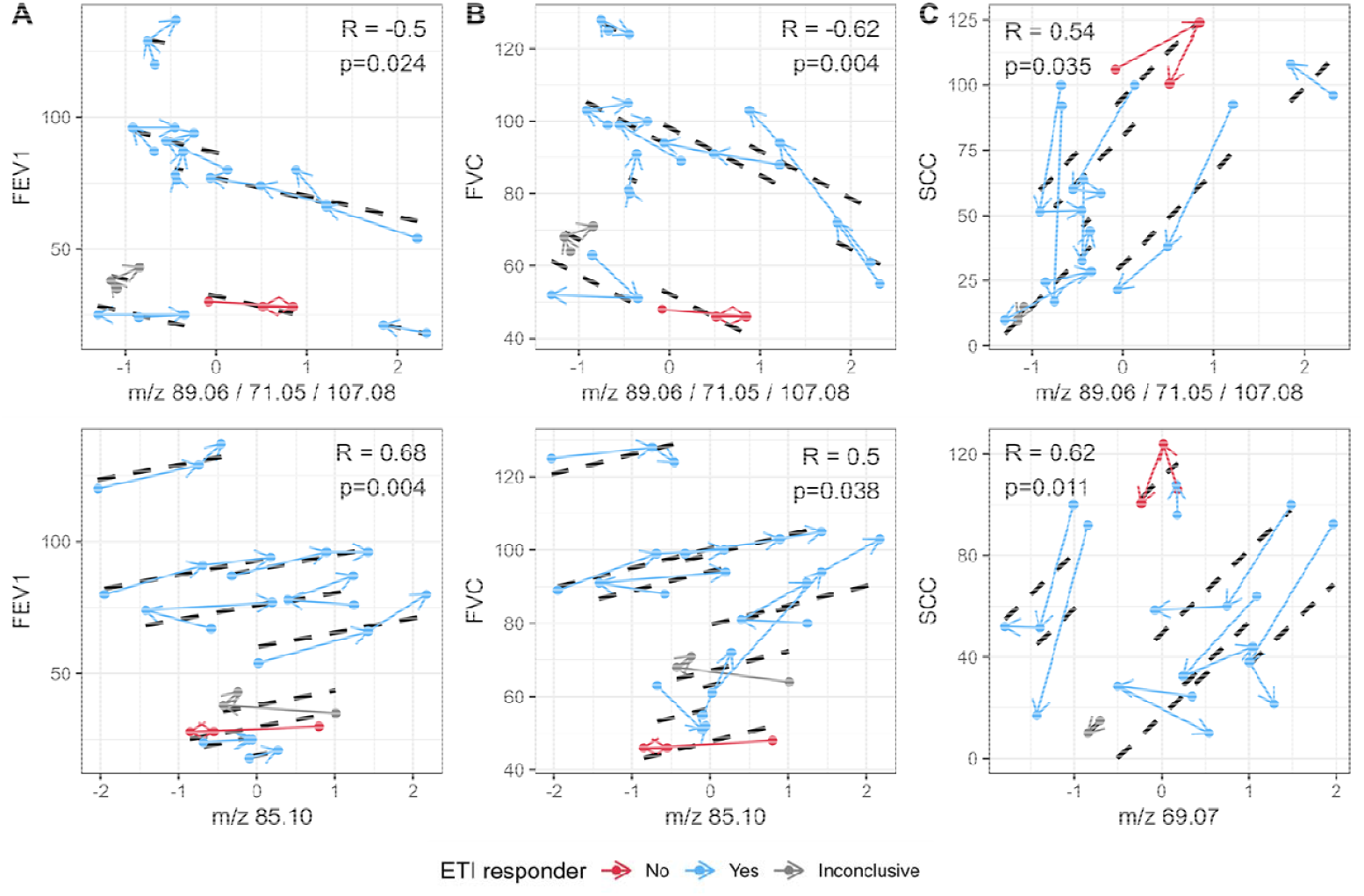
Repeated measures correlations (rmcorr) between two significant VOCs and percent predicted forced expiratory volume in 1 second (ppFEV□) (**A**), percent predicted forced vital capacity (ppFVC) (**B**), and sweat chloride concentration (SCC) (**C**). The data pertaining to each individual patient, colour-coded according to the drug response status, are linked sequentially by arrows in the order of visits (V0→V1→V2), whilst the black dotted line denotes the estimated correlation. The first row shows the same feature (*m/z* cluster [89.06/71.05/107.08]), whilst the second row shows a common feature for (**A**) and (**B**) (*m/z* 85.10), and another one (*m/z* 69.07) for (**C**). All significant correlations are shown in Supplementary Figure 7.

#### 3.2.4 Feature annotation

In addition to the five previously described clusters, supposedly corresponding to methanol, ethanol, acetic, propionic and butyric acids, several tentative annotations were proposed for the 20 features highlighted by the statistical analyses (Table 2). Features *m/z* 73.06, *m/z* 87.08 and *m/z* 135.11 were annotated as 2-butanone, 2-pentanone and o-cymene, respectively, based on an in-house library. Although structural isomers cannot be dismissed, *m/z* 69.09 is widely recognised as isoprene. Considering their exact mass and associated chemical formula, *m/z* 74.06 may correspond to N,N-dimethylformamide, propanamide or hydrated propanenitrile, *m/z* 103.08 could be pentanoic acid, whilst *m/z* 63.04 may be a water cluster of acetaldehyde. Finally, *m/z* 41.04, *m/z* 55.05, *m/z* 57.07, *m/z* 71.09, *m/z* 85.10, *m/z* 99.12 and *m/z* 139.15 most likely correspond to alkenes and/or fragments of larger compounds.

## 4 Discussion

This prospective, proof-of-concept study provides the first evidence that ETI rapidly modifies the metabolic composition of exhaled breath in adults with CF and that these short-term alterations tend to evolve towards levels observed in healthy individuals. By combining real-time PTR-MS with a multilevel statistical framework, we identified early and progressive volatilomic trajectories reflecting the biological response to ETI, and we observed significant associations between selected VOCs and clinical readouts. These findings highlight the potential of breath analysis as a non-invasive method for monitoring response to CFTR modulator therapies.

Disease- and pathogen-specific VOC signatures were previously reported in the breath of pwCF [21–23]. However, the impact of CFTR modulators has remained largely unexplored. Recent cross-sectional investigations have yielded heterogeneous results, partly because the widespread use of ETI blurs the distinction between treatment effects and disease-related metabolic signatures. Whilst Mustafina et *al* did not detect ETI-related VOCs by direct PTR-MS analysis [24], Woollam et *al* identified downregulated long-chain aldehydes using off-site gas chromatography-mass spectrometry (GC-MS) [25]. Previous work had reported some modifications induced by the dual therapy lumacaftor/ivacaftor after three months of treatment [13,26], including reductions in 2-butanone (*m/z* 73.06) and 2-pentanone (*m/z* 87.08) [26], which align with our findings on ETI. Our study further demonstrates that such changes emerge within a week of ETI initiation and continue to evolve over the first month. In a longitudinal, pilot study [27], we have previously shown early temporal response to ETI in children using GC-MS, also showing correlations between 2-butanone (*m/z* 73.06) and lung function tests. Of note, the overlap between our findings and previous GC-MS studies is limited, which most likely reflects differences in analytical technologies, populations, sampling approaches, and timepoints. Indeed, the metabolite coverage of GC-MS and PTR-MS differs substantially. PTR-MS provides rapid, high-sensitivity quantification of small, highly volatile molecules through soft ionisation, whereas GC-MS better resolves large compounds and structural isomers. These approaches should therefore be viewed as complementary. Importantly, our study is the first to include a healthy control cohort analysed on the same instrument, allowing us to contextualise ETI-induced changes relative to physiological levels.

Two distinct temporal patterns emerged from the RM-ASCA+ model. A first group of VOCs showed monotonous evolution over time, suggesting a progressive biological effect and a slow adaptation under ETI. A second group of features displayed non-monotonous trajectories, characterised by either a delayed response at one month, or an acute effect at week one followed by stabilisation or a return to baseline at one month. These divergent patterns may reflect different mechanistic layers of ETI action, possibly involving CFTR-dependent and independent mechanisms, including rapid correction of epithelial ion transport, early inflammatory shifts, changes in mucus rheology or clearance, cell membrane restructuring, and slower microbiota reshaping and metabolic rewiring.

Many of the modified VOCs (including SCFAs, ketones, aldehydes, and small alkenes) can be linked to metabolic pathways relevant to CF pathophysiology. We observed that SCFAs such as acetic (*m/z* 61.03), butyric (*m/z* 89.06) or pentanoic (*m/z* 103.08) acids were elevated in pwCF at baseline and decreased towards healthy levels after treatment. Their origin may be multifactorial, involving both pulmonary and extra-pulmonary sources. ETI has been shown to modulate the lung [28] and gut [29] microbiota, although previous studies did not report major changes in faecal SCFAs [29]. In the airways, SCFAs have been detected in sputum, where they seem to influence microbial growth and inflammatory responses [30]. Although their exact immune role is debated [31], their normalisation in breath could therefore reflect both microbiome restructuring and reduced inflammatory burden. Other VOCs, including acetaldehyde (*m/z* 63.04), ethanol (*m/z* 47.05), isoprene (*m/z* 69.09), and selected ketones, point towards energy metabolism. Acetate (*m/z* 61.03), acetaldehyde (*m/z* 63.04), and ethanol (*m/z* 47.05) are by-products of the glycolytic pathway, and their non-monotonous evolution suggests a dual acute and delayed metabolic response to ETI. Isoprene (*m/z* 69.09), derived from cholesterol biosynthesis [32], and 2-pentanone (*m/z* 87.08) were shown to rapidly rise in breath at exercise onset. The decrease of all these metabolites is consistent with a reduction in energy expenditure following ETI initiation. Finally, saturated hydrocarbons, commonly reported in breath [12], are poorly detected by PTR-MS due to weak proton affinity [33]. Several significant features may correspond to short-chain alkenes, likely arising from a mixture of endogenous origins (e.g. lipid decarboxylation [34] or membrane turnover) and analytical effects (in-source fragmentation [35]). Although identification remains putative, these metabolomic signals suggest that ETI rapidly impacts the microbiota and shifts the metabolic balance towards a more energy-efficient state. Their consistent modification in responders, unmasked by both longitudinal (LMM) and multivariate (PLS-DA) models, together with correlation with clinical outcomes, underpins the robustness of the highlighted features as potentially relevant indicators for treatment monitoring.

Using multilevel PLS-DA, a set of 11 VOCs accurately classified patient visits, achieving AUC values ≥0.84 at V0 and V1 and reaching 0.96 at V2. Moreover, several VOCs correlated with clinical outcomes which builds upon recent findings published by Woollam *et al*. and by our team [25,27]. Ten VOCs correlated with at least one clinical readout, primarily lung function. Notably, butyric acid (*m/z* 89.06) was associated with ppFEVD, ppFVC, and SCC, and it also changed significantly in responders, making it a promising candidate surrogate biomarker. These results, obtained from brief breath manoeuvres requiring minimal patient cooperation, indicate early and progressive drug-induced biological changes aligning with clinical improvement and underscore the utility of real-time breathomics for clinical monitoring. This suggests that breath analysis could detect treatment benefit even in cases where spirometry or SCC provide limited discriminatory power; for example, in individuals with preserved lung function, young children, or patients with atypical or delayed responses. As such, breathomics could contribute to more precise evaluation of CFTR modulators, especially in the context of theratyping, compassionate use, or emerging modulators targeting rare mutations.

This study has several strengths, including the use of real-time mass spectrometry, the integration of healthy controls analysed on the same instrument, and the application of complementary multilevel statistical models, which identified consistent volatilomic patterns. Some limitations must also be acknowledged. The sample size was modest as few patients were eligible for the study (most patients from our centre were already on ETI before study start). The study duration also prevented assessment of long-term stabilisation (as seen with lumacaftor/ivacaftor [13]).

In summary, this study demonstrates that ETI induces rapid and measurable changes in breath VOC profiles, that these changes tend to evolve towards healthy levels, and that they correlate with respiratory function. Real-time breath analysis therefore emerges as a promising, non-invasive tool to monitor treatment response in CF. Beyond ETI, this approach could facilitate the evaluation of new CFTR modulators, support theratyping strategies, and potentially extend to non-CF airway diseases involving CFTR dysfunction, such as COPD, asthma, or bronchiectasis. [36,37].

## Data Availability

All data generated or analysed during this study are included in this published article and its supplementary information files.

## 5 Declarations of Interest

ISG reports a Vertex Innovation Award, consulting fees and travel support from Vertex therapeutics. CR and SGD are co-founders of Exhalon. The other authors declare that they have no competing interests.

## 6 Authors Contributions

EB, HS, DG, SGD designed and planned the study; HS, SDM, DG, SGD conducted the study; CR, NH led the data analysis; EB, HS, EL, DG, SGD interpreted the results; EB, HS, SGD wrote the manuscript; ISG, PD revised the manuscript; all authors read and approved the final manuscript; SGD is responsible for the content of this study as the guarantor.

## 7 Funding

This work was supported by Région Île de France (VolatolHom, SESAME 2016 and MeLoMane, DIM 1HEALTH 2019), Fondation Foch (VolatolHom and VOC-info), and financial grants from the European Cystic Fibrosis Society and CF Europe (2020-2022) and Vaincre la Mucoviscidose (RC20220503003).

## Acknowledgments

The authors wish to acknowledge Thao Nguyen Khoa (Cochin hospital, Paris) for SCC analyses; Antoine Bertrand, Pheal company, for assistance in clinical trial promotion; Bedra Bessaidi, Beatrice d’Urso and Flavie Barret for technical investigation work. During the preparation of this work, the authors used an artificial intelligence tool solely to assist with language editing and to improve the clarity of the manuscript. The authors reviewed and validated all content and take full responsibility for the accuracy and integrity of the final submission.

## 9 Data availability statement

All data supporting the findings of this study, including summary breath metabolomics results and clinical outcomes, are included in the main manuscript and Supplementary Information. Further information about the in-house VOC library used to provide putative identification of detected VOCs can be obtained upon reasonable request to the corresponding author.

## Notes

### Clinical Trial

NCT05295524

### Funding Statement

This work was supported by Region Ile de France (VolatolHom, SESAME 2016 and MeLoMane, DIM 1HEALTH 2019), Fondation Foch (VolatolHom and VOC-info), and financial grants from the European Cystic Fibrosis Society and CF Europe (2020-2022) and Vaincre la Mucoviscidose (RC20220503003).

### Author Declarations

Patients were participating to the PHEAL-KAFTRIO study, which was approved by an ethics committee (Comite de protection des personnes Sud-Est I, 2021-A03119-32, NCT05295524). All patients provided written informed consent.

### Summary of Updates

This version of the manuscript has been revised to update the data analysis methodology.

