## Supplementary Materials for "Real-time breath metabolomics to assess early response to CFTR modulators in adults with cystic fibrosis: an open-label proof-of-concept study"

**SUPPLEMENTARY METHODS**

***PTR-Qi-TOF MS operating parameters***

A hollow cathode ion source produces reagent ions (H₃O⁺) and protons are transferred from H₃O⁺ to VOCs in a drift tube forming [VOC+H]⁺ ions based on proton affinity. A quadrupole then conveys and focuses the protonated analytes towards the TOF analyser, which separates them according to their mass-over-charge ratio (*m/z*). Operating parameters were as follows: electric field strength (E/N) 127.8 Td; source voltage 120 V; drift tube pressure, temperature and voltage were set to 3.8 mbar, 60°C and 959 V, respectively. Mass spectra were acquired up to *m/z* 392 with a time resolution of 1 s; experimental mass resolution was approximately 5,000 (m/Δm).

***Data pre-processing***

Data were pre-processed using the ptairMS R package [1]. Mass axis calibration was performed using the following reference ions: *m/z* 21.02, 29.01, 60.05, 203.94, and 330.85. Estimated mean resolution of the instrument across calibration peaks was 5,000 [range: 3,000–8,000] (m/Δm). A peak deconvolution algorithm enabled the separation of isobaric peaks with a minimum *m/z* difference of 0.01 Da (Figure S1). The ptairMS workflow then estimates the evolution of each detected peak along the time axis to provide ion traces. Exhalation phase detection was based on the trace of the acetone isotope (*m/z* 60.05). Each feature was then quantified by the mean intensity measured across the five exhalation phases, expressed in count per second.

Cystic fibrosis (CF) patient samples were aligned with a cohort of healthy young adults using Gaussian kernel density estimation. The standard deviation of the smoothing kernel was set to 30 ppm, i.e. the maximum allowable deviation between acquisitions on the *m/z* axis. Features detected in at least 70% of CF samples were retained for further analysis. Missing values, representing ions not detected during individual sample peak detection, were imputed by integrating the noise at the exact missing *m/z* in the raw data.

The cps data were normalised using the probabilistic quotient normalisation (PQN) algorithm [2,3], ensuring that signal intensities were corrected for unwanted variation from non-biological sources. Then, peaks significantly higher in exhalation phases than in the background were identified through a *t-*test analysis (*p*-value <1e-10); and features significant in at least 20% of the CF samples were retained.

Saturated ions (e.g., acetone, H_3_O⁺, H_2_O–H_3_O⁺) and isotopic peaks were excluded. Hierarchical clustering analysis (HCA) was used to identify clusters amongst the remaining features, employing Pearson correlation as the distance metric. We made the hypothesis that clusters with within-group correlations greater than 0.9 originated from a unique parent molecule. The *m/z* difference between such features were calculated to identify addition/loss of H_2_O ($\Delta m/z 18.02)$. When the parent molecule of a cluster was chemically identified, only the parent ion intensity was retained.

***Statistical analysis - Univariate linear mixed model (LMM)***

We used the following linear mixed-effect model for each feature:

$$y_{ij}^{k}=\beta_{0}+\beta_{1}V1_{ij}+ \beta_{2}V2_{ij}+{b_{0}}_{i}+\epsilon_{ij}$$

where:

$y_{ij}^{k}$ the PQN-normalised, standardised value of the feature $k$intensity, for the patient $i$ at time level $j$ $\in$ [V0,V1,V2]

$$V1_{ij}=1 if j=V1, 0 else and V2_{ij}=1 if j=V2, 0 else$$

$\beta_{1}$ and $\beta_{2}$ represent the effect of time at V1 and V2 compared to V0 (baseline); and $\beta_{0}$ the level at baseline.

${b_{0}}_{i}$ the random effect for the patient $i$

The F-test (lmerTest package [4] using Satterthwaite's method) is used to test if

H0: $({\beta_{1,}\beta}_{2})=0$

H1: $({\beta_{1,}\beta}_{2}) \neq0$

$(\beta_{0},\beta_{1},\beta_{2})$ are the three coefficients representing the fixed effect for each feature used for the repeated measures ANOVA–simultaneous component analysis plus (RM-ASCA+) model [5,6].

***Statistical analysis - RM-ASCA+, PLS-DA and RFE***

A repeated measures ANOVA–simultaneous component analysis plus (RM-ASCA+) was performed, consisting of a principal component analysis (PCA) applied to the LMM fixed-effect coefficients of each feature. This approach enables visualisation of the main temporal evolution patterns in the data, and quantification of the proportion of variance explained by time by averaging the marginal R² of the LMM models.

A PCA was performed to identify the main source of variation, and a multilevel partial least-squares – discriminant analysis (PLS-DA) model was applied using the mixOmics R package [7,8], with visits as the predictive features and individuals as the multilevel factor. The model's performance was evaluated using a leave-one-out cross-validation procedure at the patient level (i.e. eight-fold), along with the area under the curve (AUC) of the receiver operating characteristic (ROC) for each visit, with 95% confidence interval bootstrap.

A recursive feature elimination (RFE) algorithm was used to identify the features contributing most to the classification model's performance. Within each cross-validation fold, features were ranked according to variable importance in projection (VIP) metrics, and the top *n* features were retained to rebuild the model and test it on the held-out patients. Results were aggregated across folds. Values of *n* between 5 and 30 were tested; the optimal number was chosen based on the average cross-validation AUC across the three visits.

***Feature annotation***

Tentative chemical identification was based primarily on exact mass match with the human breathomics database [9] and likelihood of compound ionisation by proton transfer reaction [10].

Some annotations were refined using others’ published work [11–13] as well as an in-house library of volatile organic compounds (VOC). The latter integrated features detected by proton-transfer reaction - mass spectrometry (PTR-MS) that correlated (r > 0.6) with peaks identified by thermal desorption - 2D-gas chromatography – mass spectrometry in corresponding CF and healthy breath samples (BIO-CFTR, NCT02965326 and VOC-COMPARE, NCT06020521, respectively).

**SUPPLEMENTARY TABLES**

**Supplementary Table 1.** List of concomitant treatments and bronchial exacerbations during the study.

| **Patient** | **Chronic antimicrobial drugs** | **Acute respiratory infections during the study and associated prescribed drugs** |
| --- | --- | --- |
| 01 | Azithromycin, oral  Tobramycin and aztreonam, inhaled | COVID-19, treatment with amoxicillin and ciprofloxacin |
| 02 | Azithromycin, oral | - |
| 03 | - | - |
| 04 | - | - |
| 05 | Azithromycin, oral  Pristinamycin and amoxicillin-clavulanic acid, oral | - |
| 06 | Azithromycin, oral | - |
| 07 | Azithromycin, oral  Tobramycin and colomycin, inhaled | COVID-19, treatment with ceftazidime and doxycycline |
| 08 | Azithromycin, oral | - |
| 09 | - | - |
| 10 | Azithromycin, oral | Viral syndrome |

**Supplementary Table 2.** List of detected *m/z* features with tentative annotations and *p*-values from the LMM and correlation analyses.

| Detected *m/z* | Formula | Possible annotations | *p*-value LMM | *p*-value correlation ppFEV_1_ | *p*-value correlation ppFVC | *p*-value correlation SCC |
| --- | --- | --- | --- | --- | --- | --- |
| 28.01 |  |  | 0.121 | 1 | 1 | 0.069 |
| 31.00 |  |  | 0.172 | 1 | 0.649 | 1 |
| 31.02 | [CH_2_O+H]^+^ | formaldehyde | 1 | 0.553 | 1 | 1 |
| 33.03/ 51.04 | [CH_4_O+H]^+^ | methanol | 0.091 | 0.448 | 1 | 0.210 |
| 41.04 | [C_3_H_4_+H]^+^ | allene/ prop-1-yne | 0.154 | 0.199 | 1 | 0.595 |
| 41.05 |  |  | 1 | 1 | 1 | 1 |
| 42.03 | [C_2_H_3_N+H]^+^ | acetonitrile | 0.568 | 0.226 | 0.019 | 1 |
| 43.99 |  |  | 0.156 | 0.469 | 0.232 | 1 |
| 45.00 | [CO_2_+H]^+^ |  | 0.172 | 0.331 | 0.304 | 0.272 |
| 47.05/ 65.06 | [C_2_H_6_O+H]^+^ | ethanol/ methoxymethane | 0.073 | 0.163 | 1 | 1 |
| 49.01 | [CH_4_S+H]^+^ | methanethiol | 1 | 0.446 | 0.457 | 0.259 |
| 53.04 | [C_4_H_4_+H]^+^ | but-1-en-3-yne | 0.313 | 1 | 1 | 1 |
| 55.04 |  |  | 0.489 | 0.211 | 0.868 | 0.766 |
| 55.05 | [C_4_H_6_+H]^+^ | buta-1,3-diene/ but-1-yne/ but-2-yne | **0.041** | 1 | 1 | 1 |
| 57.07 | [C_4_H_8_+H]^+^ | but-1-ene/ (E)-but-2-ene/ (Z)-but-2-ene/ cyclobutane/ 2-methylprop-1-ene | **0.029** | 0.503 | **0.050** | 0.724 |
| 58.04 |  |  | 0.765 | 1 | 0.499 | 1 |
| 60.05 |  |  | 0.507 | 0.753 | 1 | 0.701 |
| 60.07 |  |  | 0.114 | 1 | 0.766 | 1 |
| 61.03/ 43.02/ 79.04 | [C_2_H_4_O_2_+H]^+^ | acetic acid/ 2-hydroxyacetaldehyde/ methyl formate | **0.052** | 0.992 | 0.147 | 0.095 |
| 63.01 |  |  | 0.743 | 1 | 1 | 0.345 |
| 63.03 | [C_2_H_6_S+H]^+^ | methylsulfanylmethane/ ethanethiol | 0.612 | 1 | 1 | 0.490 |
| 63.04 |  |  | **0.027** | 0.206 | 1 | 1 |
| 67.05 | [C_5_H_6_+H]^+^ | cyclopenta-1,3-diene/ 2-methylbut-1-en-3-yne/ pent-3-en-1-yne/ (E)-pent-3-en-1-yne | 0.542 | 1 | 0.522 | 1 |
| 67.07 |  |  | **0.016** | 0.641 | 1 | 0.220 |
| 68.06 |  |  | 0.642 | 0.632 | 1 | 0.115 |
| 69.07 | [C_5_H_8_+H]^+^ | isoprene/ (3E)-penta-1,3-diene/ (3Z)-penta-1,3-diene/ penta-1,4-diene/ pent-2-yne | 0.060 | 0.106 | 1 | 0.078 |
| 69.09 |  |  | 0.116 | 0.526 | 1 | **0.011** |
| 71.09 | [C_5_H_10_+H]^+^ | cyclopentane/ 2-methylbut-2-ene/ 3-methylbut-1-ene/ 2-methylbut-1-ene/ (E)-pent-2-ene/ (Z)-pent-2-ene | 0.200 | 0.976 | **0.004** | 1 |
| 71.11 |  |  | 0.255 | 1 | 0.169 | 0.158 |
| 73.03 | [C_3_H_4_O_2_+H]^+^ | propanedial/ 2-oxopropanal | 1 | 0.545 | 0.826 | 0.369 |
| 73.06 | [C_4_H_8_O+H]^+^ | butanal/ butan-2-one/ 2-methylpropanal/ vinyloxyethane | 0.222 | 0.182 | **0.015** | 0.583 |
| 74.06 | [C_3_H_7_NO+H]^+^ | N,N-dimethylformamide/ propanamide | **0.038** | **0.029** | 0.364 | 0.260 |
| 75.04/ 57.03 | [C_3_H_6_O_2_+H]^+^ | methyl acetate/ 1,3-dioxolane/ 1-hydroxypropan-2-one/ ethyl formate/ 2-methoxyacetaldehyde/ propionic acid | 0.413 | 0.739 | 0.289 | 0.440 |
| 77.06 | [C_3_H_8_O_2_+H]^+^ | dimethoxymethane/ 2-methoxyethanol/ propane-1,2-diol | 0.268 | 0.220 | 0.520 | 0.454 |
| 77.09 |  |  | 0.105 | 0.692 | 1 | 0.325 |
| 79.05 | [C_6_H_6_+H]^+^ | benzene | 0.230 | 1 | 1 | 0.597 |
| 79.08 |  |  | 0.065 | 0.331 | 1 | 0.399 |
| 80.05 | [C_5_H_5_N+H]^+^ | pyridine | 0.476 | 0.868 | 0.753 | 1 |
| 81.02 |  |  | 0.721 | 1 | 1 | 1 |
| 81.07 | [C_6_H_8_+H]^+^ | cyclohexa-1,3-diene/ cyclohexa-1,4-diene/ (3Z)-hexa-1,3,5-triene/ 5-methylcyclopenta-1,3-diene/ 1-methylcyclopenta-1,3-diene/ 4-methylenecyclopentene/ 3-methylenecyclopentene/ (Z)-3-methylpent-3-en-1-yne | 0.237 | 1 | 0.975 | 0.660 |
| 85.03 | [C_4_H_4_O_2_+H]^+^ | 2H-furan-5-one | 0.367 | 1 | 1 | 1 |
| 85.06 | [C_5_H_8_O+H]^+^ | NA/cyclopentanone/ 3,4-dihydro-2H-pyran/ 3-methylbut-2-enal/ (E)-2-methylbut-2-enal/ 3-methylbut-3-en-2-one/ pent-1-en-3-one/ (E)-pent-3-en-2-one | 0.697 | 1 | 0.779 | 0.708 |
| 85.10 | [C_6_H_12_+H]^+^ | cyclohexane/ 3,3-dimethylbut-1-ene/ 2,3-dimethylbut-1-ene/ 2,3-dimethylbut-2-ene/ 3-methylenepentane/ hex-1-ene/ (E)-hex-2-ene/ (Z)-hex-2-ene/ (E)-hex-3-ene/ methylcyclopentane/ 2-methylpent-1-ene/ 3-methylpent-1-ene/ 4-methylpent-1-ene/ 2-methylpent-2-ene/ (E)-3-methylpent-2-ene/ (E)-4-methylpent-2-ene/ (Z)-4-methylpent-2-ene | **0.001** | **0.004** | **0.038** | 0.102 |
| 85.13 |  |  | 0.803 | 0.989 | 1 | 1 |
| 87.04 | [C_4_H_6_O_2_+H]^+^ | butane-2,3-dione/ (E)-but-2-enoic acid/ tetrahydrofuran-2-one/ 2,3-dihydro-1,4-dioxine | 0.332 | 0.057 | 1 | 0.730 |
| 87.08 | [C_5_H_10_O+H]^+^ | 3-methylbutanal/ 2-methylbutanal/ 3-methylbutan-2-one/ pentanal/ pentan-2-one | **0.006** | 0.325 | 1 | 0.514 |
| 88.05 |  |  | 1 | 1 | 1 | 1 |
| 88.08 | [C_4_H_9_NO+H]^+^ | 2-methylpropanamide | 0.770 | 0.779 | 1 | 1 |
| 89.06/ 71.05/ 107.08 | [C_4_H_8_O_2_+H]^+^ | 2-methylpropanoic acid/ ethyl acetate/ butyric acid/ 1,4-dioxane/ 3-hydroxybutanal/ propyl formate/ methyl propanoate | **0.051** | **0.024** | **0.004** | **0.035** |
| 91.06 | [C_4_H_10_S+H]^+^ | 1-methylsulfanylpropane | 0.763 | 1 | 1 | 0.706 |
| 93.04 | [C_2_H_8_O_2_Si+H]^+^ | dihydroxy(dimethyl)silane | 0.939 | 0.260 | 0.199 | 1 |
| 93.07 | [C_7_H_8_+H]^+^ | toluene | 0.180 | 1 | 1 | 0.840 |
| 95.01 | [C_2_H_6_O_2_S+H]^+^ | methylsulfonylmethane | 0.178 | 1 | 0.201 | 0.243 |
| 95.05 | [C_6_H_6_O+H]^+^ | hydron;phenoxide/ 2-vinylfuran | 0.061 | 1 | 1 | 1 |
| 95.09 | [C_7_H_10_+H]^+^ | cyclohepta-1,3-diene/ 1,2-dimethylcyclopenta-1,3-diene/ 5,5-dimethylcyclopenta-1,3-diene/ hepta-1,3,5-triene/ 1-methylcyclohexa-1,4-diene/ 3-methylenecyclohexene | 0.165 | 1 | 1 | 1 |
| 99.04 | [C_5_H_6_O_2_+H]^+^ | 2-furylmethanol/ 2-methyl-2H-furan-5-one/ 4-methyl-2H-furan-5-one | 0.598 | 0.419 | 1 | 0.962 |
| 99.12 | [C_7_H_14_+H]^+^ | cycloheptane/ 1,1-dimethylcyclopentane/ 1,2-dimethylcyclopentane/ 1,3-dimethylcyclopentane/ 2,4-dimethylpent-2-ene/ (E)-3,4-dimethylpent-2-ene/ 4,4-dimethylpent-1-ene/ hept-1-ene/ (E)-hept-2-ene/ (E)-hept-3-ene/ 3-methylhex-1-ene/ 4-methylhex-1-ene/ 5-methylhex-1-ene/ 2-methylhex-2-ene/ (E)-3-methylhex-2-ene/ (E)-4-methylhex-2-ene/ (Z)-2-methylhex-3-ene /(E)-2-methylhex-3-ene/ (Z)-3-methylhex-3-ene/ (E)-5-methylhex-2-ene | 0.097 | **0.037** | 0.068 | 1 |
| 103.04 | [C_4_H_6_O_3_+H]^+^ | acetyl acetate | 0.884 | 1 | 1 | 0.660 |
| 103.08 | [C_5_H_10_O_2_+H]^+^ | propyl acetate/ isopropyl acetate/ 3-methylbutanoic acid/ pentanoic acid/ ethyl propanoate | **0.052** | **0.009** | 0.281 | 0.132 |
| 105.03 |  |  | 0.199 | 0.062 | 0.293 | 0.382 |
| 105.07 | [C_8_H_8_+H]^+^ | styrene | 0.562 | 1 | 1 | 1 |
| 108.96 |  |  | 0.599 | 0.512 | 1 | 1 |
| 109.07 | [C_7_H_8_O+H]^+^ | phenylmethanol/ m-cresol/ p-cresol/ 2-allylfuran | 0.239 | 1 | 0.509 | 1 |
| 111.05 | [C_6_H_6_O_2_+H]^+^ | 1-(2-furyl)ethanone/ 5-methylfuran-2-carbaldehyde | 0.683 | 0.672 | 0.796 | 0.663 |
| 111.08 | [C_7_H_10_O+H]^+^ | 1-methoxycyclohexa-1,3-diene/ 2-ethyl-5-methyl-furan/ 3-methylcyclohex-2-en-1-one/ 2,3,5-trimethylfuran | 0.528 | 0.609 | 1 | 1 |
| 113.06 |  |  | 0.138 | 1 | 0.987 | 1 |
| 113.13 | [C_8_H_16_+H]^+^ | cyclooctane/ 1,1-dimethylcyclohexane/ 1,2-dimethylcyclohexane/ 1,3-dimethylcyclohexane/ 1,4-dimethylcyclohexane/ 3,4-dimethylhex-1-ene/ 2,5-dimethylhex-1-ene/ 5,5-dimethylhex-1-ene/ 2,5-dimethylhex-2-ene/ 3,5-dimethylhex-2-ene/ (E)-2,5-dimethylhex-3-ene/ 1-ethyl-2-methyl-cyclopentane/ 2-methyl-4-methylene-hexane/ methylcycloheptane/ 3-methyleneheptane/ 2-methylhept-1-ene/ 4-methylhept-1-ene/ 6-methylhept-1-ene/ 2-methylhept-2-ene/ (E)-3-methylhept-2-ene/ (E)-4-methylhept-2-ene/ 6-methylhept-2-ene/ (E)-5-methylhept-2-ene/ oct-1-ene/ (E)-oct-2-ene/ (E)-oct-3-ene/ (E)-oct-4-ene/ (1R,3R)-1,2,3-trimethylcyclopentane/ 1,2,3-trimethylcyclopentane/ 1,2,4-trimethylcyclopentane/ 2,4,4-trimethylpent-2-ene/ 2,3,4-trimethylpent-2-ene/ (E)-3,4,4-trimethylpent-2-ene | 1 | 0.914 | 1 | 0.630 |
| 115.08 | [C_6_H_10_O_2_+H]^+^ | 2-methylallyl acetate/ hexane-2,5-dione/ (E)-4-methylpent-2-enoic acid/ propyl prop-2-enoate | 0.816 | 1 | 1 | 0.661 |
| 117.09 | [C_6_H_12_O_2_+H]^+^ | butyl acetate/ isobutyl acetate/ ethyl butanoate/ hexanoic acid/ 4-methylpentanoic acid/ propyl propanoate/ methyl 2-methylbutanoate | 0.458 | 0.659 | 0.426 | 0.441 |
| 118.07 | [C_8_H_7_N+H]^+^ | 5H-cyclopenta[b]pyridine | 0.674 | 0.494 | 0.685 | 0.948 |
| 119.09 | [C_9_H_10_+H]^+^ | allylbenzene/ indane/ 1-methyl-2-vinyl-benzene/ isopropenylbenzene/ 1-methyl-4-vinyl-benzene/ [(E)-prop-1-enyl]benzene | 0.276 | 0.063 | 0.225 | 0.761 |
| 123.04 | [C_7_H_6_O_2_+H]^+^ | hydron;benzoate | 0.431 | 0.833 | 0.679 | 1 |
| 125.96 |  |  | 0.636 | 1 | 1 | 0.415 |
| 126.01 |  |  | 0.911 | 0.866 | 1 | 1 |
| 126.97 |  |  | 0.880 | 1 | 0.839 | 0.268 |
| 127.15 | [C_9_H_18_+H]^+^ | 1-ethyl-3-methyl-cyclohexane/ 1-ethyl-4-methyl-cyclohexane/ (E)-non-2-ene/ (E)-non-3-ene/ (Z)-non-3-ene/ 1,2,3-trimethylcyclohexane/ 1,2,4-trimethylcyclohexane/ 1,1,2-trimethylcyclohexane/ (1S,3S)-1,2,3-trimethylcyclohexane/ 1-methylpentylcyclopropane | 0.799 | 1 | 1 | 1 |
| 129.13 | [C_8_H_16_O+H]^+^ | 2-ethylhexanal/ 3-methylheptan-2-one/ 6-methylheptan-2-one/ 6-methylheptan-3-one/ octanal/ octan-2-one/ octan-3-one/ 1-ethylcyclohexanol/ 3-methylheptan-4-one | 0.273 | 1 | 1 | 0.846 |
| 135.11 | [C_10_H_14_+H]^+^ | 1-isopropyl-2-methyl-benzene/ butylbenzene/ 1-isopropyl-4-methyl-benzene/ 1-methyl-4-propyl-benzene/ 1,4-diethylbenzene/ 1,3-diethylbenzene/ 1-ethyl-2,3-dimethyl-benzene/ 1-ethyl-3,5-dimethyl-benzene/ 2-ethyl-1,4-dimethyl-benzene/ 1-ethyl-2,4-dimethyl-benzene/ 4-ethyl-1,2-dimethyl-benzene/ 5-isopropenyl-2-methyl-cyclohexa-1,3-diene/ 1-isopropyl-3-methyl-benzene/ 1-methyl-2-propyl-benzene/ 1,2,3,4-tetramethylbenzene/ 1,2,3,5-tetramethylbenzene/ 1,2,4,5-tetramethylbenzene | 0.495 | 1 | **0.043** | 0.373 |
| 137.13 | [C_10_H_16_+H]^+^ | 2,2-dimethyl-3-methylene-norbornane/ (3E)-3,7-dimethylocta-1,3,6-triene/ (3E,5E)-3,7-dimethylocta-1,3,5-triene/ 4-isopropenyl-1-methyl-cyclohexene/ 7-methyl-3-methylene-octa-1,6-diene/ 3-isopropyl-6-methylene-cyclohexene/ 5-isopropyl-2-methyl-cyclohexa-1,3-diene/ 6,6-dimethyl-2-methylene-norpinane/ 2,6,6-trimethylbicyclo[3.1.1]hept-2-ene/ 1-isopropyl-4-methyl-cyclohexa-1,3-diene/ 1-isopropyl-4-methylene-cyclohexene/ 1-isopropyl-4-methyl-cyclohexa-1,4-diene/ 4-isopropylidene-1-methyl-cyclohexene/ 3,7,7-trimethylbicyclo[4.1.0]hept-3-ene/ 1,7,7-trimethylbicyclo[2.2.1]hept-2-ene | 0.135 | 0.237 | 0.976 | 0.354 |
| 139.08 | [C_8_H_10_O_2_+H]^+^ | 2-phenoxyethanol | 0.150 | 0.604 | 1 | 0.242 |
| 139.15 | [C_10_H_18_+H]^+^ | 2,7-dimethylocta-1,6-diene/ (6E)-2,6-dimethylocta-2,6-diene/ (6Z)-2,6-dimethylocta-2,6-diene/ 2,7-dimethylocta-2,6-diene/ 1-isopropenyl-4-methyl-cyclohexane/ 3-isopropyl-6-methyl-cyclohexene/ (3R,6S)-3-isopropyl-6-methyl-cyclohexene/ 1-isopropyl-4-methyl-cyclohexene/ 2-methylprop-1-enylcyclohexane/ 3,3,6-trimethylhepta-1,5-diene/ 3,3,5-trimethylhepta-1,5-diene | **0.027** | 0.956 | 0.810 | 1 |
| 145.12 | [C_8_H_16_O_2_+H]^+^ | octanoic acid/ 2-ethylhexanoic acid | 0.412 | 0.664 | 0.560 | 0.232 |
| 163.13 | [C_8_H_18_O_3_+H]^+^ | 1-(2-butoxyethoxy)ethanol | 1 | 1 | 1 | 0.476 |
| 163.20 |  |  | 0.886 | 1 | 1 | 0.167 |
| 177.16 | [C_13_H_20_+H]^+^ | heptylbenzene | 1 | 0.746 | 1 | 0.184 |
| 205.14 |  |  | 0.205 | 1 | 0.485 | 0.245 |
| 205.20 |  |  | 1 | 0.761 | 0.620 | 1 |
| 207.18 | [C_14_H_22_O+H]^+^ | 2,5-ditert-butylphenol/ 4-(1,1,3,3-tetramethylbutyl)phenol/ 2,4-ditert-butylphenol | 0.369 | 1 | 1 | 1 |
| 227.16 |  |  | 0.659 | 1 | 1 | 0.357 |
| 235.21 |  |  | 0.379 | 0.970 | 1 | 1 |
| 247.23 |  |  | 0.526 | 1 | 0.567 | 0.370 |
| 371.10 |  |  | 0.061 | 0.869 | 1 | 1 |

LMM: linear mixed-effects model; ppFEV_1_: percent predicted forced expiratory volume in 1 second; ppFVC: percent predicted forced vital capacity; SCC: sweat chloride concentration; significant *p*-values (<0.05) after false discovery rate (FDR) correction are in bold.

**SUPPLEMENTARY FIGURES**

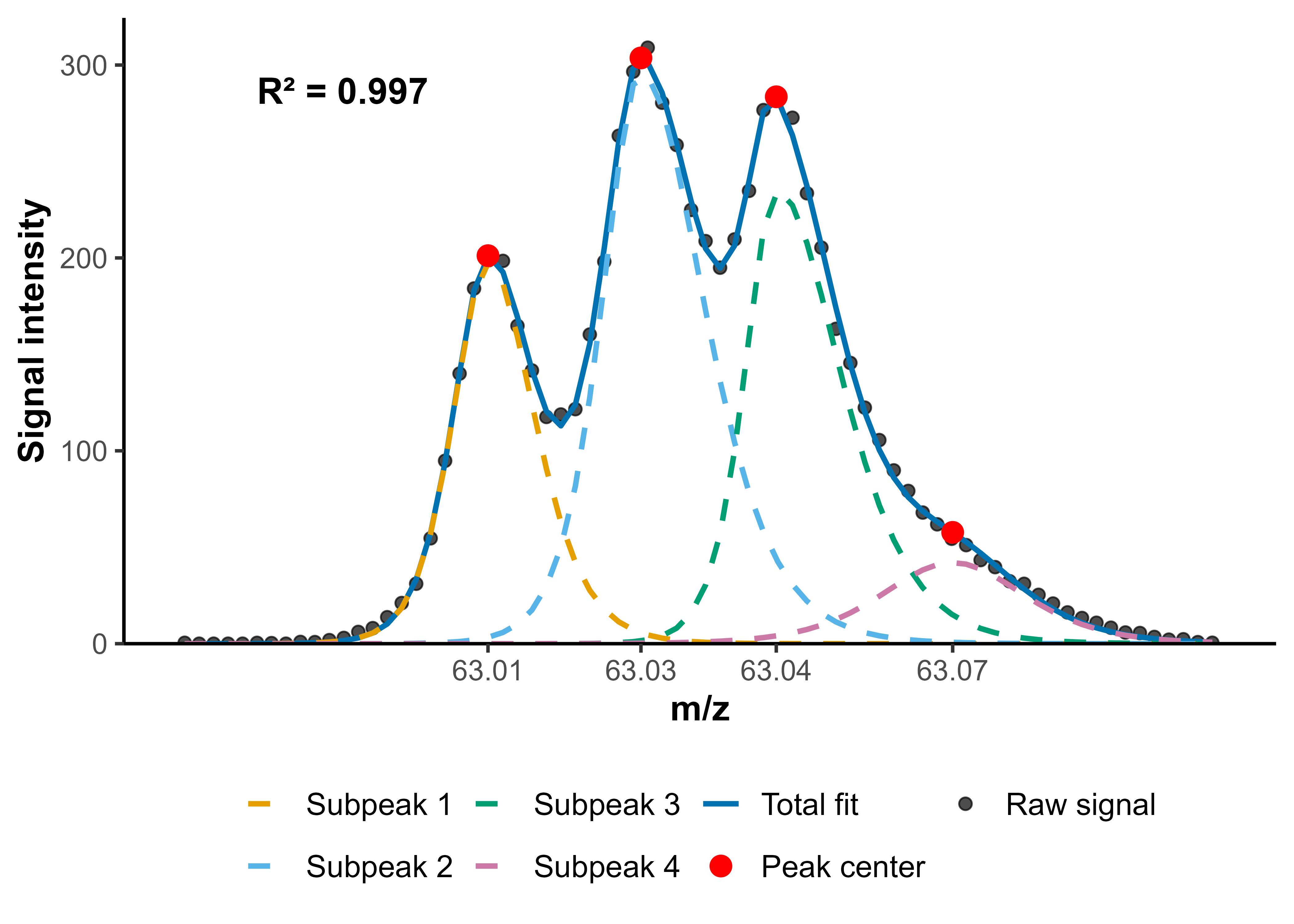

**Supplementary Figure 1**. Example of peak deconvolution with the software ptairMS around *m/z* 63, showing the possible separation of peaks with an *m/z* difference of 0.01 Da.

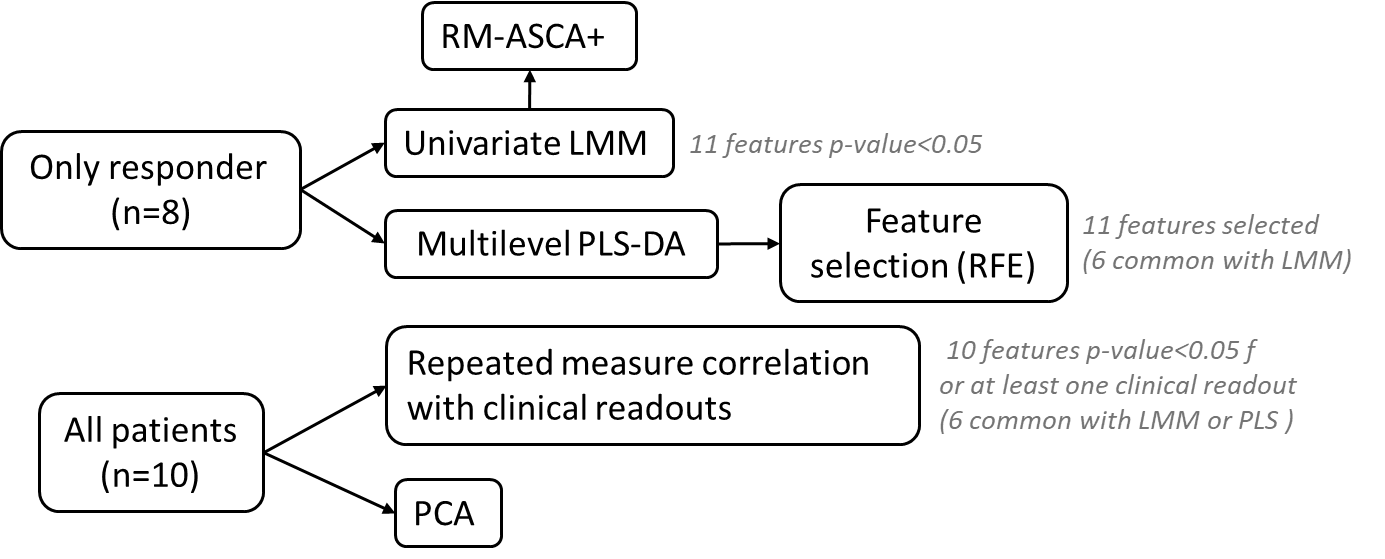

**Supplementary Figure 2**. Statistical analysis workflow.

RM-ASCA+: repeated measures ANOVA–simultaneous component analysis plus; LMM: linear mixed-effects model; PLS-DA: partial least-squares – discriminant analysis; RFE: recursive feature elimination; PCA: principal component analysis.

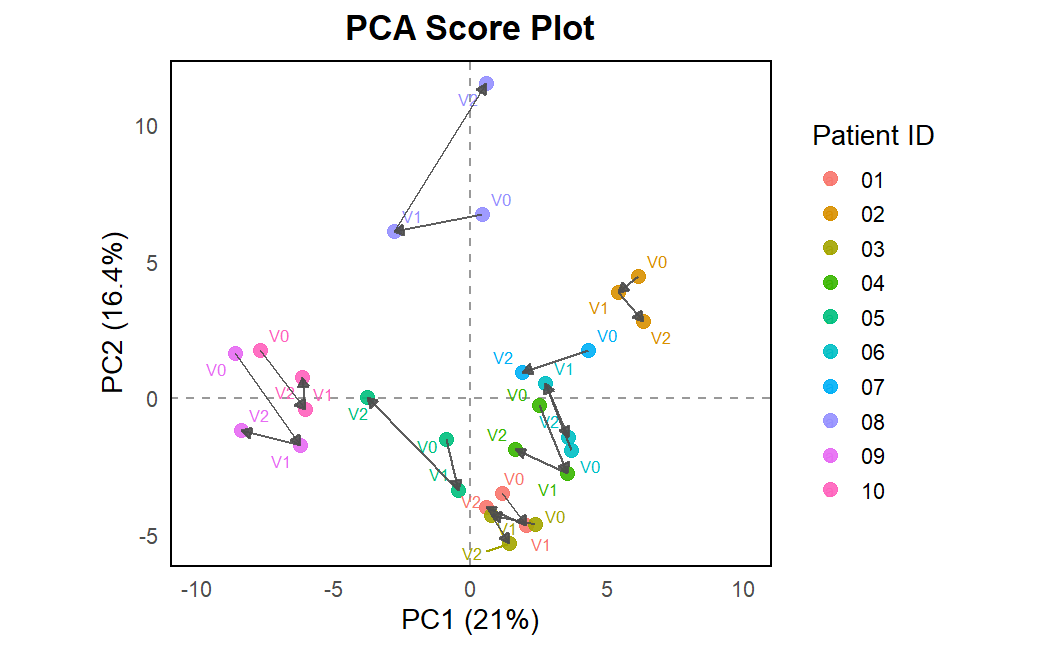

**Supplementary Figure 3**. Principal component analysis (PCA) of breath samples collected from each patient at baseline (V0), after one week (V1), and after one month of ETI treatment (V2). Data points are colour-coded by patient, with points connected in chronological order of visits.

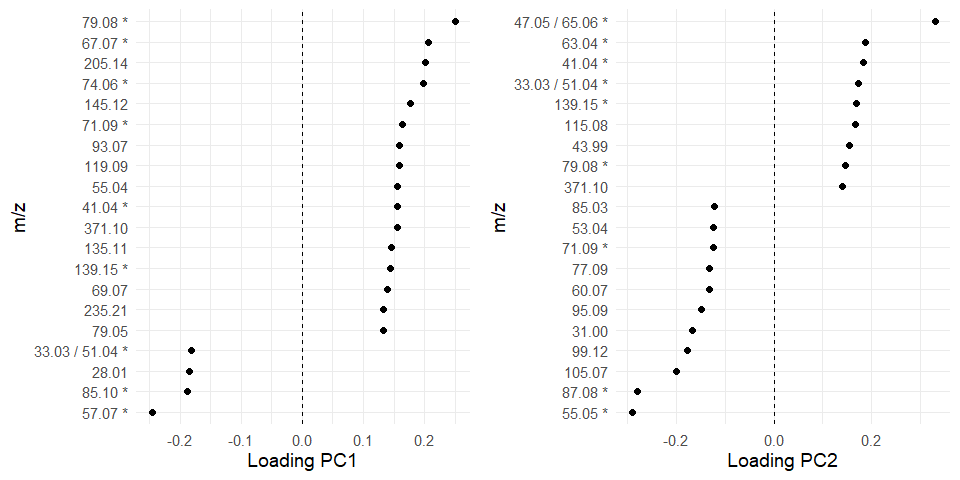

**Supplementary Figure 4**. The twenty most influential loadings for the two principal components (PCs) of the repeated measures ANOVA–simultaneous component analysis plus (RM-ASCA+) model. Features selected for the multilevel partial least-squares – discriminant analysis (PLS-DA) model are indicated with an asterisk (*). Negative loading values denote contributions in the opposite direction relative to the corresponding PC.

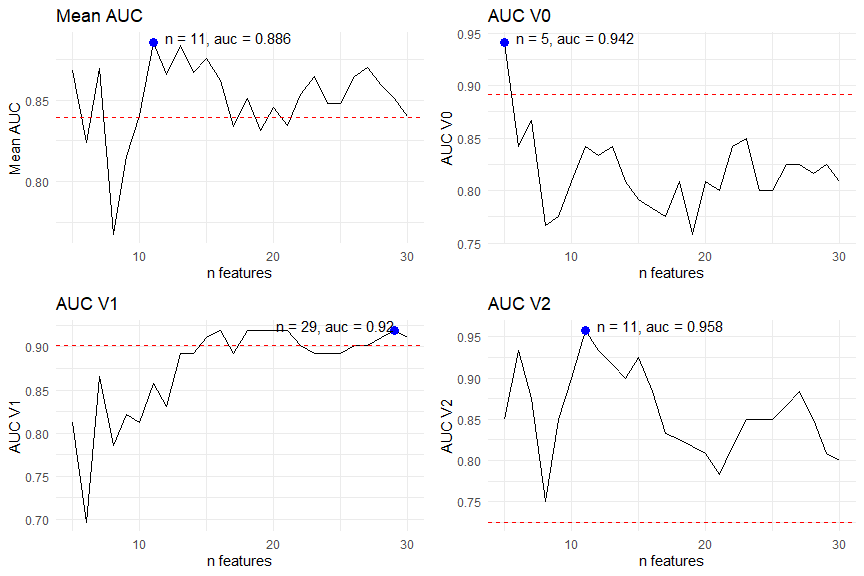

**Supplementary Figure 5.** Area under the curve (AUC) as a function of the number of selected features using the recursive feature elimination (RFE) algorithm. The top left panel shows the mean AUC across the three visits. The top right, and bottom panels correspond to baseline (V0), one week (V1), and one month (V2). The red line sets the AUC value of the model using all features. The optimal number of features (*n*) was selected based on the highest mean AUC (*n*=11).

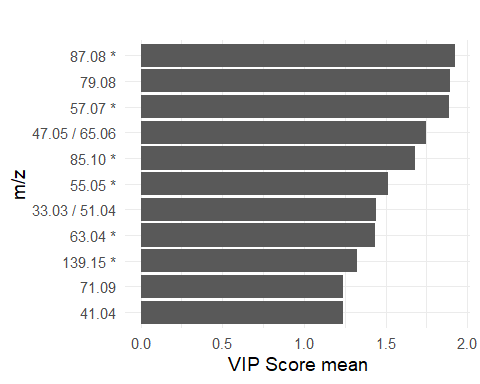

**Supplementary Figure 6.** Mean variable importance in projection (VIP) scores of the 11 features selected by recursive feature elimination (RFE), resulting from the cross-validation of the multilevel partial least-squares – discriminant analysis (PLS-DA) model. Features selected through the linear mixed-effect models (LMM) are indicated with an asterisk (*).

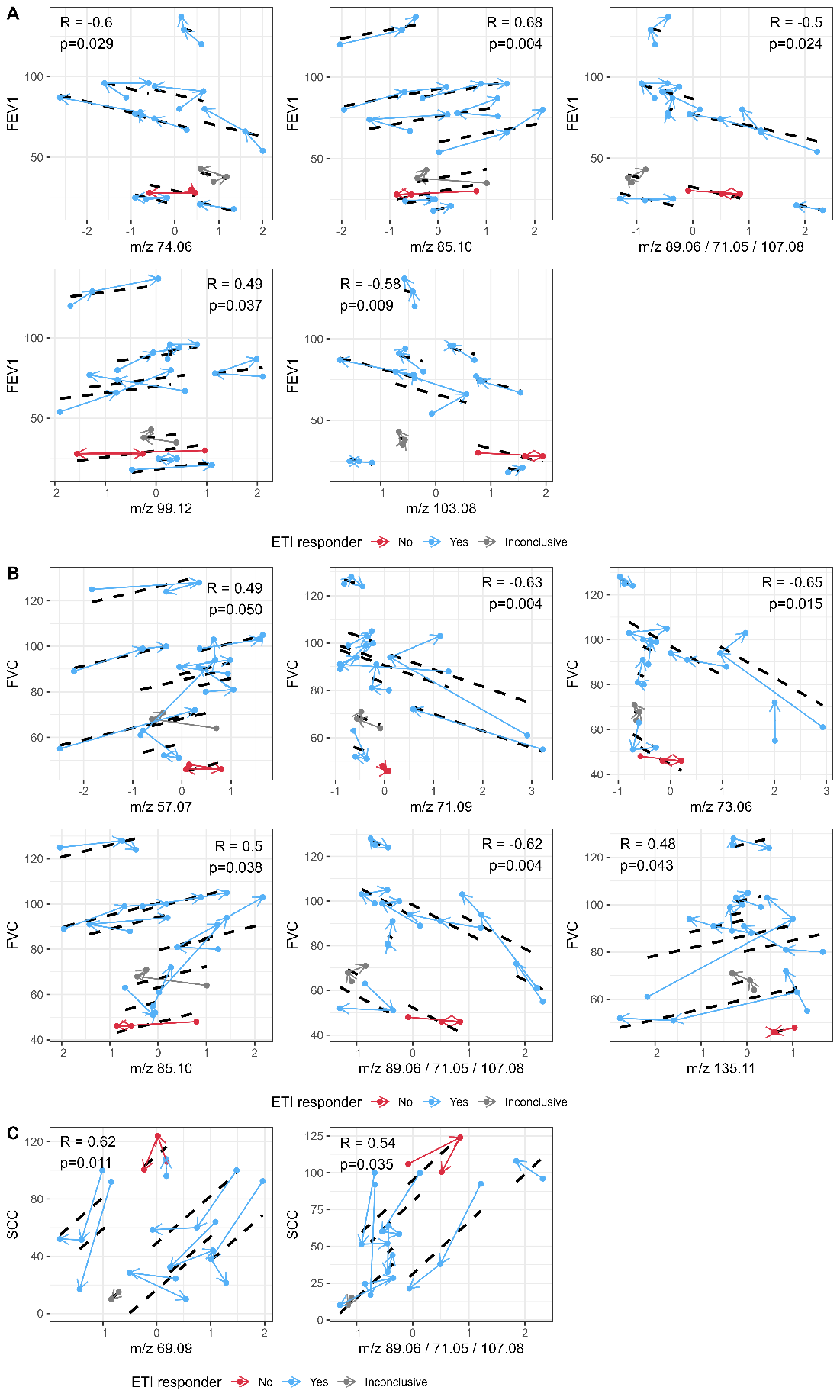

**Supplementary Figure 7**. Correlations between exhaled breath features and (**A**) percent predicted forced expiratory volume in 1°second (ppFEV₁), (**B**) percent predicted forced vital capacity (ppFVC), and (**C**) sweat chloride concentration (SCC), assessed using a repeated measures correlation test (rmcorr) [14]**.** The data pertaining to each individual patient, colour-coded according to the drug response status, are linked sequentially by arrows in the order of clinical visits (V0 → V1 → V2), whilst the black dotted line denotes the estimated correlation.
